# Multicellular immune ecotypes within solid tumors predict real-world therapeutic benefits with immune checkpoint inhibitors

**DOI:** 10.1101/2024.07.19.24310726

**Authors:** Xuefeng Wang, Tingyi Li, Islam Eljilany, Vineeth Sukrithan, Aakrosh Ratan, Martin McCarter, John Carpten, Howard Colman, Alexandra P. Ikeguchi, Igor Puzanov, Susanne Arnold, Michelle Churchman, Patrick Hwu, Paulo C. Rodriguez, William S. Dalton, George J. Weiner, Ahmad A. Tarhini

**Affiliations:** Department of Biostatistics and Bioinformatics, H. Lee Moffitt Cancer Center and Research Institute, Tampa, FL 33612, USA; Departments of Cutaneous Oncology and Immunology, H. Lee Moffitt Cancer Center and Research Institute, Tampa, FL 33612, USA; Department of Internal Medicine, Ohio State University and Arthur G James Comprehensive Cancer Center, Columbus, OH 43210 USA; Center for Public Health Genomics, School of Medicine, University of Virginia, Charlottesville, VA 22903, USA; Department of Surgery, University of Colorado Cancer Center, Aurora, CO 80045, USA; City of Hope Comprehensive Cancer Center, Duarte, CA 91010, USA; Department of Neurosurgery, School of Medicine, University of Utah, Salt Lake City, UT 84132, USA; University of Texas MD Anderson Cancer Center, Houston, TX 77030, USA.; Department of Medicine, Roswell Park Comprehensive Cancer Center, Buffalo, NY 14263, USA; Department of Medical Oncology, University of Kentucky Markey Cancer Center, Lexington, KY 40536, USA; Aster Insights, Hudson, FL 34667, USA; H. Lee Moffitt Cancer Center and Research Institute, Tampa, FL 33612, USA.; Department of Immunology, H. Lee Moffitt Cancer Center and Research Institute, Tampa, FL 33612, USA; Department of Internal Medicine, Carver College of Medicine, University of Iowa Health Care, Iowa City, IA 52242, USA

**Keywords:** Cell Atlas, Cell State, Cell Type, Ecotype, Immunotherapy, Melanoma

## Abstract

**Background:** Cancer initiation, progression, and immune evasion depend on the tumor microenvironment (TME). Thus, understanding the TME immune architecture is essential for understanding tumor metastasis and therapy response. This study aimed to create an immune cell states (CSs) atlas using bulk RNA-seq data enriched by eco-type analyses to resolve the complex immune architectures in the TME.

**Methods:** We employed EcoTyper, a machine-learning (ML) framework, to study the real-world prognostic significance of immune CSs and multicellular ecosystems, utilizing molecular data from 1,610 patients with multiple malignancies who underwent immune checkpoint inhibitor (ICI) therapy within the ORIEN Avatar cohort, a well-annotated real-world dataset.

**Results:** Our analysis revealed consistent ICI-specific prognostic TME carcinoma ecotypes (CEs) (including CE1, CE9, CE10) across our pan-cancer dataset, where CE1 being more lymphocyte-deficient and CE10 being more proinflammatory. Also, the analysis of specific immune CSs across different cancers showed consistent CD8+ and CD4+ T cell CS distribution patterns. Furthermore, survival analysis of the ORIEN ICI cohort demonstrated that ecotype CE9 is associated with the most favorable survival outcomes, while CE2 is linked to the least favorable outcomes. Notably, the melanoma-specific prognostic EcoTyper model confirmed that lower predicted risk scores are associated with improved survival and better response to immunotherapy. Finally, de novo discovery of ecotypes in the ORIEN ICI dataset identified Ecotype E3 as significantly associated with poorer survival outcomes.

**Conclusion:** Our findings offer important insights into refining the patient selection process for immunotherapy in real-world practice and guiding the creation of novel therapeutic strategies to target specific ecotypes within the TME.

**Trial Registration:** NCT04526730

## INTRODUCTION

Cancer is a leading cause of mortality globally, accounting for nearly 10 million deaths in 2020.^1^ Fortunately, in recent years, immune checkpoint inhibitors (ICIs) have emerged as transformative agents in cancer therapy, modulating costimulatory immune checkpoints to restore T cell effector functions and anti-cancer immunity, and have demonstrated significantly favorable outcomes in treating cancer.^2,3^ However, responses to ICIs are variable by tumor type and within the same histologies. One of the valid reasons for this is that uncontrolled cellular proliferation is a hallmark of malignant tumors that manifest a complex ecosystem of countless interactions between tumor cells and host immune cells within a complex tumor microenvironment (TME).^4–6^ This warrants deeper insights into the tumor-immune dynamics within the TME to predict responsiveness, allowing for personalized therapeutic strategies and identifying resistance mechanisms that may guide future drug development.^7^

The TME is pivotal in cancer initiation, progression, and immune evasion.^8,9^ This dynamic system harbors a rich and diverse ecosystem of immune cells, fibroblasts, and endothelial cells, among others, intricately interacting with cancer cells ^10,11^. Hence, understanding the immune infrastructure within the TME becomes critical to elucidate the mechanisms governing tumor metastasis and response to therapies, including ICIs^12^. Moreover, an in-depth investigation of the underlying immune cell states (CSs) through a detailed atlas can uncover such predictive markers and complex mechanisms in patients treated with ICIs. ^2,13–16^

Undoubtedly, the advent of RNA sequencing (RNA-seq) technologies has revolutionized the landscape of cancer research, offering exceptional insights into the molecular signatures and complex pathways active within tumors.^17^ Besides, advances in computational deconvolution methods have allowed the inference of the abundance of different cell types within the TME utilizing bulk RNA-seq data.^18^ As a leading example, CIBERSORTx has proven great applicability in immunogenomics, allowing the imputation of gene expression profiles and estimation of the abundance of member cell types in complex tissues using gene expression data. Subsequently, innovative techniques have enabled the estimation of gene expression levels unique to individual cell types, commonly called purification approaches, unveiling distinct biological states within a homogeneous cell population characterized by their unique transcriptome patterns.^19,20^ Eco-type analyses utilizing the EcoTyper program allow a superior understanding of tumor cellular heterogeneity. Each eco-type is defined as a frequent group of cell types in conjunction with a specific combination of CSs.^21^ EcoTyper was developed as an extension of CIBERSORTx to identify and quantify CSs through deconvolving TME using large-scale bulk transcriptomics data, scRNA-seq, or spatial transcriptomics. This deconvolution process aims to generate cell types and states that complement the ones obtained by single-cell studies. ^22,23^ It has been shown to enhance the prognostic accuracy for tumor progression and patient prognosis across diverse tumor types, drawing from many data sources. ^24^

This study sought to construct an immune CS atlas leveraging bulk RNA-seq data enriched through eco-type analyses to resolve the complex immune architectures within the TME, distinguish the distinct immune CSs, and foster a deeper understanding of the correlations between immune cell states and responsiveness to ICIs. The outcomes of this study provide insights into the underlying mechanisms impacting response and resistance to ICIs.

## METHODS

### Patient Cohort and Data Compilation

This research incorporated a retrospective examination of clinical data and gene expression profiles from consenting patients collected through Total Cancer Care® (TCC.) Protocol (NCT03977402) and the Avatar® project conducted within the Oncology Research Information Exchange Network (ORIEN), which includes 18 collaborating cancer centers participating in TCC ^25,26^ Subjects participating in TCC provided written informed consent to allow the use of their tumor and blood biospecimens for genomic and transcriptomic analyses and corresponding clinical data as part of the standard clinical practice to manage their disease. The ORIEN Avatar cohort in our study consisted of 14,997 individuals, of whom 1,610 patients were treated with ICIs and constituted our target population. The ICI medication group included “Ipilimumab,” “Nivolumab,” “Dostarlimab,” “Pembrolizumab,” “Avelumab,” “Atezolizumab,” “Cemiplimab,” and “Durvalumab.” The study was conducted in accordance with the ethical standards of the Declaration of Helsinki, along with approval by the Institutional Review Board (IRB) at each participating institution (Advara IRB # Pro00014441). The supplementary materials provide additional information on the ICI cohorts used for validation

### RNA Sequencing

The procedure for RNA sequencing for the ORIEN Avatar project was conducted according to methodologies outlined in a white paper previously released (https://www.asterinsights.com/white-paper/renal-cell-carcinoma-rwd-data/). The necessary data on RNA expression were sourced from the ORIEN database, necessitating the download of multiple FASTQ files for further examination.

### Quantification of RNA Gene Expression

Quantifying gene expression in our study involved multiple technical steps. The initial phase employed Bbduk software (version 38.96) to remove adapter sequences from RNA-seq reads, as detailed at “https://sourceforge.net/projects/bbmap/.”^27^ This was followed by aligning the trimmed reads to the human reference genome (CRCH38/hg38) using STAR software (version 2.7.3a), accessible at “https://github.com/alxdobin/STAR.”^28^ The integrity of the RNA samples was evaluated using the RNA-Seq Quality Control (RNA-SeQC) software (version 2.3.2), found at “https://github.com/getzlab/rnaseqc.” ^29^ The computation of gene expression levels was performed using the Transcripts Per Million (TPM) metric, following alignment with the GeneCode build version 32 reference annotation through the RNA-Seq by Expectation Maximization (RSEM) software (version 1.3.1), available at “https://github.com/deweylab/RSEM.”^30^ For the purposes of analysis, TPM values were transformed to a logarithmic scale (Log2 [TPM+1]) after an increment of +1 for linear conversion. Any batch effects were adjusted using the ComBat method in the sva package (version 3.34.0), documented at “https://doi.org/doi:10.18129/B9.bioc.sva.”^31^

### Ecotyper Framework

EcoTyper was conducted to deconvolute the RNA-seq data from the ORIEN ICI cohort and investigate CSs and ecotypes. Using our bulk RNA-seq data, cell states were first identified and categorized within each cell type, which is identified in the original discovery pan-cancer TCGA dataset by Luca et al.^23^ Subsequently, the algorithm establishes CS co-occurrence models to define cellular communities referred to as ecosystems. In this study, we focused on assessing the prognostic significance of 10 carcinoma ecosystems (CEs) that have been previously defined. The identified prognostic CEs in the ORIEN melanoma cohort were then independently validated in external cohorts of patients with melanoma treated with ICI.^32–35^

### Predictive Model Development

Our analyses initially aimed to retrospectively evaluate the clinical associations of predicted risk scores based on the ecotype-based model with patients’ overall survival (OS) in melanoma. Consequently, a model was constructed and trained using data from 159 patients with melanoma enrolled in the ORIEN dataset. Initially, a univariable Cox regression analysis was conducted to explore the individual prognostic significance of various ecological features (eco-groups). Subsequently, a more comprehensive analysis was undertaken using a 10-fold elastic-net Cox regression model with an α parameter set to 0.3. Furthermore, an Adaptive Lasso approach was employed to refine the model further and enhance interpretability. To begin with, a 10-fold L2 penalized regression was conducted, yielding coefficient estimates penalized with lambda set to the one standard error (SE) criterion (lambda = 1SE). Afterwards, a 5-fold L1 penalized Cox regression (lasso) was performed, utilizing α set to 0.4.

Additionally, the penalization weights applied during this step were calculated based on the absolute values of the coefficients obtained from the previous L2 penalized regression, effectively incorporating ridge-derived information into the penalty structure. Moreover, a partially penalized Cox model was explored to investigate the impact of targeted regularization. Specifically, CE groups CE1, CE2, CE9, and CE10 were fixed, while the remaining eco-types were penalized using α set to 0.4. Finally, a stepwise selection procedure based on the Akaike Information Criterion (AIC) was employed to refine the model architecture further.

### Statistical Analysis

Our statistical analysis plan involved quite a few steps. Firstly, it began with a univariable Cox regression analysis to assess the individual prognostic significance of eco-groups. The associations between each eco-group and OS as survival outcomes were quantified using Cox regression coefficients and corresponding *p*-values.

Secondly, multivariate Cox regression analysis was conducted to assess the independent prognostic value of the selected eco-groups while adjusting for potential confounders. A stepwise variable selection procedure based on AIC was employed to identify the subset of eco-groups that collectively provided the best predictive performance. Model assumptions, including the proportional hazards assumption, were evaluated using Schoenfeld residuals and log-minus-log plots.

Thirdly, regularization techniques such as LASSO and ridge regression were utilized to prevent overfitting. Cross-validation was employed to select the optimal regularization parameter (lambda) and assess model performance. The resulting penalized regression models were used to identify the most influential eco-groups in predicting survival outcomes. Fourthly, model performance was quantitatively assessed using standard survival analysis metrics, including Harrell’s concordance index (C-index). Confidence intervals (Cis) and hypothesis testing were used to assess the statistical significance of model improvements compared to baseline or competing models.

Finally, cross-validation techniques, such as k-fold cross-validation, were employed to estimate the generalization error of the prognostic model and optimize model hyperparameters. Stratified sampling was used to ensure a balanced representation of outcome events (e.g., deaths) across cross-validation folds. All statistical analyses were performed using R version 4.1.3 and relevant packages: survival, glmnet, and stepreg.

## RESULTS

### Patients Characteristics of ORIEN ICI cohort

In a comprehensive examination of the clinical characteristics of participants at baseline, our analysis identified a real-world cohort of 14,997 in the ORIEN database, among them 1610 patients treated with ICIs in the ORIEN dataset. This cohort is categorized as follows: melanoma (n=161), head and neck squamous cell carcinoma (H&N) (n= 240), non-small cell lung cancer (NSCLC) (n=270), bladder cancer (n=151), and kidney cancer (n=311) as presented in Table 1 and Supplementary Figure 1. The mean ± standard deviation (SD) age across the cohort was 61.53 (12.37) years.

**Table 1.** Baseline clinical characteristics of patients included in the ORIEN ICI cohort.

|  | **Overall\***<br>**(N=1610)** | **Melanoma**<br>**(N=161)** | **H&N**<br>**(N=240)** | **NSCLC**<br>**(N=270)** | **Others\***<br>**(N=942)** |
| --- | --- | --- | --- | --- | --- |
| **Age at study** |  |  |  |  |  |
| Mean<br>(SD) | 61.53<br>(±12.37) | 59.44<br>(±14.30) | 61.19<br>(±10.82) | 63.51<br>(± 9.58) | 61.39<br>(±13.00) |
| **Gender, n (%)** |  |  |  |  |  |
| Women | 38.70 | 38.75 | 17.92 | 46.67 | 41.72 |
| Men | 61.30 | 61.25 | 82.08 | 53.33 | 58.28 |
| **Ethnicity, n (%)** |  |  |  |  |  |
| Hispanic | 4.72 | 8.08 | 2.92 | 2.92 | 5.73 |
| Non-Hispanic | 93.42 | 90.06 | 96.25 | 96.25 | 91.93 |
| Unknown | 1.86 | 1.86 | 0.83 | 0.83 | 2.34 |
| **Tumor origin, n (%)** |  |  |  |  |  |
| Primary | 65.53 | 35.40 | 60.80 | 60.00 | 73.35 |
| Metastasis | 30.12 | 63.35 | 31.67 | 35.56 | 22.40 |
| Both | 4.35 | 1.24 | 7.50 | 4.44 | 4.25 |
| **Cancer Stage, n (%)** |  |  |  |  |  |
| Stage 0 | 0.19 | 0.00 | 0.00 | 0.00 | 0.32 |
| Stage I | 6.40 | 3.11 | 5.00 | 12.22 | 5.63 |
| Stage II | 8.20 | 9.32 | 2.92 | 14.81 | 7.54 |
| Stage III | 15.84 | 14.90 | 7.50 | 14.33 | 18.47 |
| Stage IV | 17.33 | 10.56 | 24.17 | 18.52 | 16.45 |
| Unknown | 51.99 | 62.11 | 60.42 | 40.00 | 51.59 |
| **Immuno-oncology therapy, n (%)** |  |  |  |  |  |
| Ipilimumab | 3.11 | 21.74 | 0.42 | 0.00 | 1.49 |
| Nivolumab | 26.02 | 22.36 | 27.08 | 21.11 | 27.71 |
| Pembrolizumab | 49.01 | 39.75 | 67.08 | 58.52 | 43.31 |
| Avelumab | 1.30 | 0.00 | 0.00 | 0.37 | 2.12 |
| Atezolizumab | 6.34 | 0.621 | 1.67 | 7.41 | 8.17 |
| Cemiplimab | 0.56 | 0.00 | 0.83 | 0.00 | 0.74 |
| Durvalumab | 2.55 | 0.00 | 0.83 | 10.37 | 1.17 |
| Ipilimumab+Nivolumab | 10.99 | 14.29 | 2.08 | 2.22 | 15.29 |
| Ipilimumab+Nivolumab/Pembrolizumab | 0.06 | 0.621 | 0.00 | 0.00 | 0.00 |
| Nivolumab-Relatimab-rmbw | 0.06 | 0.621 | 0.00 | 0.00 | 0.00 |

\*Others; All cancer types except melanoma. ¥Some patients have more than one cancer type.
Abbreviations: ICI; immune checkpoint inhibitor, H&N; head and neck, NSCLC; non-small cell lung cancer, SD; standard deviation.

Specifically, melanoma patients presented a slightly younger demographic with a mean ± SD age of 59.44 ± 14.30 years at the time of data collection, while NSCLC patients were the oldest with a mean ± SD age of 63.51 ± 9.58 years. Men and non-Hispanic participants were predominant (61.30% and 93.42%, respectively) across the cohort in all cancer types. The analysis of tumor origin highlighted a noteworthy prevalence of primary tumors in all cancer tumors, except for a prominent 63.35% of melanoma cases that were identified as metastatic.

The data shows potential gaps in cancer staging information or that some participants were selected regardless of their stages at the time of initial diagnosis since over half (51.99%) had stages listed as unknown. However, all subjects included who had advanced metastatic stages necessitating treatment with ICIs. Therefore, various treatments were reported across the cohort, with pembrolizumab being the most administered drug (49.01%). As expected, melanoma patients received ipilimumab at a significantly higher rate (21.74%) compared to other cancer groups.

### Global distribution of identified cell-states and multicellular ecotypes

Turning to CSs and multicellular ecotypes in the analysis of the discovery cohort, the heatmap in Figure 1A shows the distribution of multicellular communities or carcinoma ecotypes (CEs) across all samples within this real-world ORIEN ICI cohort, which comprises 1610 patients with 1732 RNAseq samples. Of these, 817 patients (77%) were successfully assigned to specific ecotypes.

**Figure 1.**
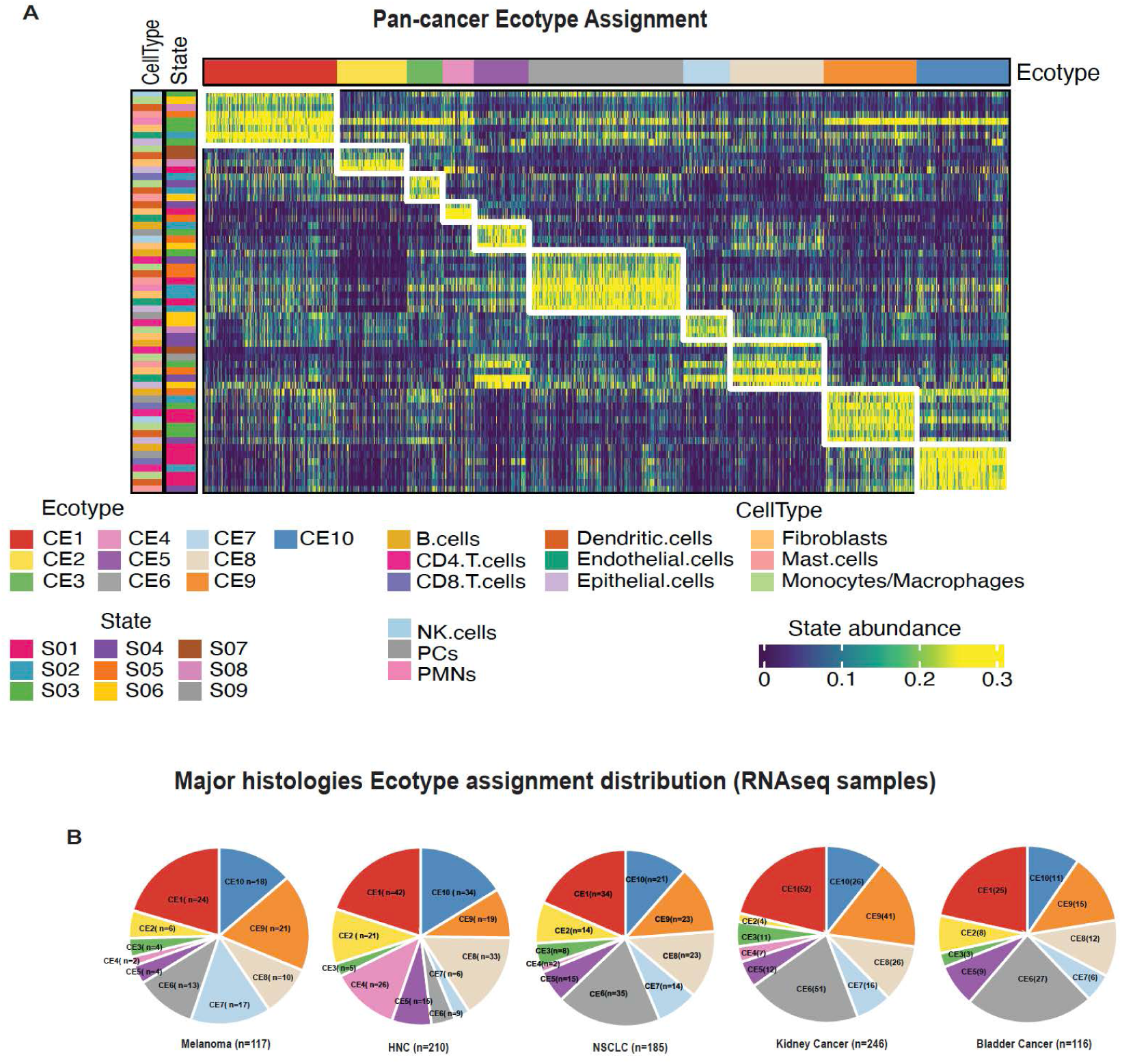
Distribution of Carcinoma Ecotypes (CEs) in the Pan-Cancer ORIEN IO Cohort. **(A)** This heatmap displays the abundance profiles of cell states and carcinoma ecotypes identified across all cancer types in the ORIEN IO cohort. Only samples assigned to pre-defined CEs (CE1-CE10) are included. Each column in the heatmap corresponds to RNA sequencing samples, and each row shows the expression of cell state marker genes within a recovered ecotype. (**B**) Pie charts illustrate the distribution of the ten CEs within five specific cancer groups: melanoma, head and neck cancer (H&N), non-small cell lung cancer (NSCLC), kidney cancer, and bladder cancer. Each segment indicates the number of assigned patients per ecotype group—abbreviations: CE, carcinoma ecotype; S, cell state.

These assignments covered all ten previously defined ecotypes across carcinomas (CE1-10).^23^ A notable trend was observed, where CE1 tumors represent more lymphocyte-deficient, and CE10 tumors represent more proinflammatory CEs. The ecotypes with the highest number of tumors assigned were CE1 and CE6 (a non-neoplastic tissue enriched cell subtype), followed by CE8 through CE10. This distribution showed slight differences from the findings in the TCGA pan-cancer discovery cohort (n=4729), where CE1 and CE8 were the most prevalent.^23^

A comparison of the distribution of ecotypes results across the five major cancer types in the cohort as illustrated in Figure 1B: melanoma, H&N, NSCLC, kidney cancer, and bladder carcinoma revealed that 80% of H&N patients, 75% of NSCLC patients, 75% of kidney cancer patients, 72% of patients with bladder cancer, and 70% of melanoma patients were successfully assigned to a pre-defined ecotype. In each cancer type, the previously known pan-cancer prognostic ecotypes CE1-CE2 and CE9-CE10 were found in around or more than half of the patients, indicating the translational potential of these four ecotypes for predicting ICI outcomes. Among these five cancer types, H&N exhibited the most distinct cancer-specific distribution, with a higher number of samples assigned to CE4 and notably fewer in CE6. This pattern aligns with the discovery cohort, where a higher prevalence of CE4, potentially linked to myogenesis, was noted in both H&N and prostate cancer (and in older male patients).

### Distribution of specific immune cell states across cancer types

As shown in Figure 2, we further investigated the distribution of specific immune CSs identified from the gene expression data from the discovery dataset and in melanoma, H&N, NSCLC, and bladder cancer. These analyses focused on different cell types and their potential role in the immune system. Looking at Figure 2A, it is apparent that the distribution of the three identified CD8 T CSs, CD4-S01 (naive/central memory T cells), CD4-S02 (late-stage effectors), and CD4-S03 (exhausted/effector memory), is consistent across the four cancer types examined. Note that CD4-S01 and CD4-S03 are key components of the two proinflammatory communities CE10 and CE9, respectively.

**Figure 2.**
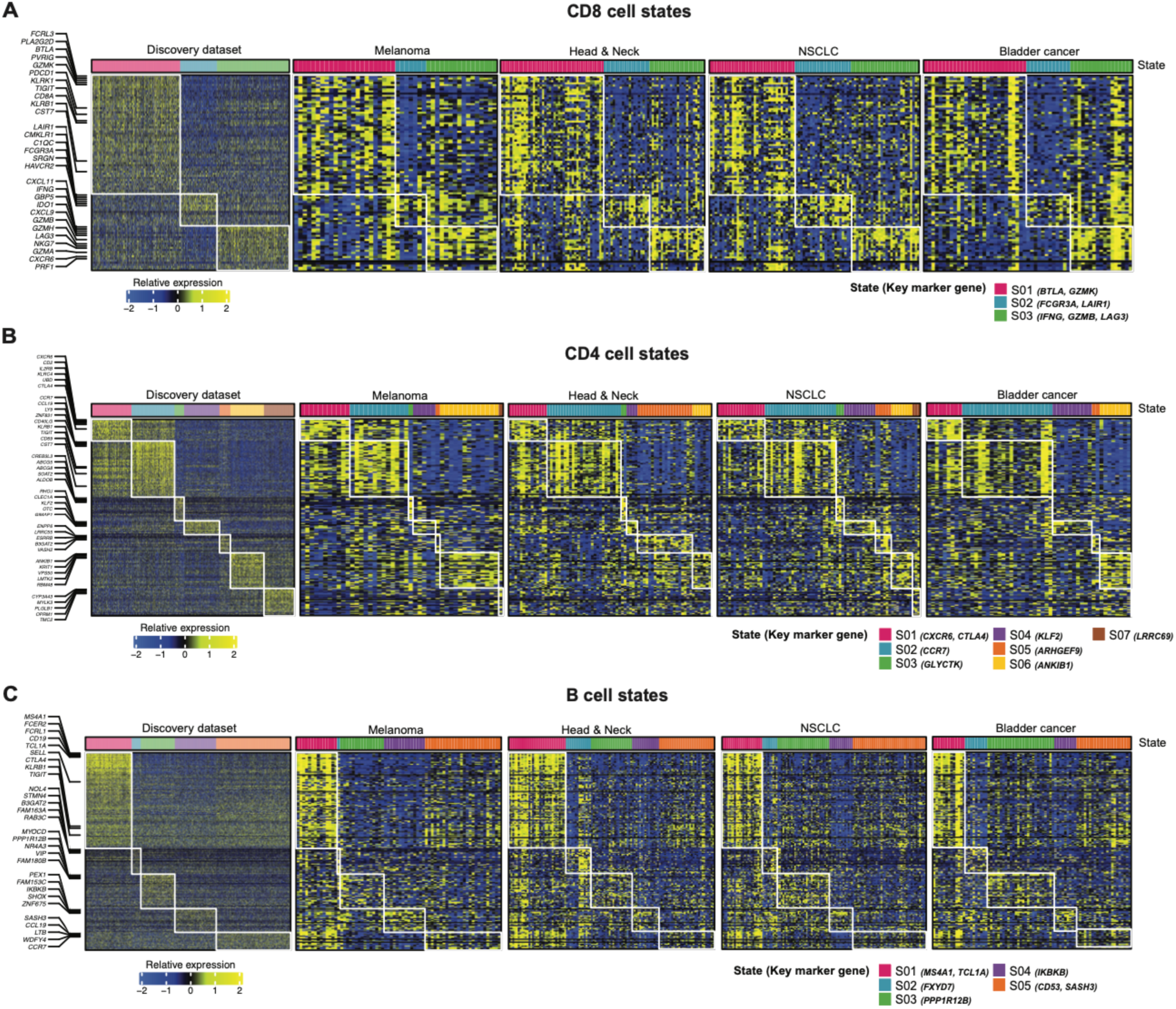
Cell State Expression Across Major Cancer Types. The figure presents heatmaps displaying detailed expression profiles of tumor-associated cell states, focusing on CD4 (**A**), CD8 (**B**), and B cells (**C**). The first heatmap in each panel details the expression of marker genes for each cell state, using data from TCGA datasets as a reference. Subsequent heatmaps illustrate the expression profiles in each major cancer type. Each row corresponds to a cell-state marker gene, and each column represents RNA sequencing samples.

In the CD4 cell type (Figure 2B), however, only the distributions of the two cell states CD4-S01 (exhausted/effector memory) and CD4-S02 (naive/central memory) remain consistent across the four cancers, accounting for half of the patients with an assigned CD4 CS assigned. These two states define the two proinflammatory ecotypes. It is observed that CD4-S05 was more enriched in H&N cancer, while CD4-S06 is more enriched in melanoma, with prominence in the specific immune environments of these cancers. Overall, NSCLC displayed a broader mixture of CD4 states, specifically S03-S06. This diversity is presumably attributed to the histological heterogeneity within NSCLC, including adenocarcinoma and squamous cell subtypes.

Regarding the distribution of B cell states, as portrayed in Figure 2C, relatively similar patterns are observed across the cancers, with certain exceptions; there was a higher prevalence of CD4-S05 (activated B cells) in melanoma and NSCLC and a relatively higher occurrence of CD4-S03 (normal-enriched) in bladder cancer.

Additionally, in the analysis of dendritic cells shown in Supplementary Figure 2, DC-S01 (myeloid cDC1) was prevalent across all four cancers, especially in melanoma and H&N. Meanwhile, DC-S05, identified as mature/normal-enriched, was more abundant in bladder cancer and NSCLC. These findings, especially the observed variations in non-CD8 immune cells, emphasize the significance of accounting for the complete range of immune CSs and interactions beyond the conventional categories of immune hot and immune cold.

### Prognostic ecotypes for ICI cohort

The clinical significance of carcinoma CS co-occurrence networks, or CEs, has been previously established by the high concordance between the identified ecotypes and prognosis across various solid tumor types.^24^ Notably, CE9, characterized by IFN-γ signaling and highly active anti-tumor immune activity, demonstrated potential in predicting ICI response in metastatic melanoma trial data.^23^ As a primary objective of this study, we conducted a thorough correlative analysis of treatment outcomes concerning all CEs within the ORIEN ICI cohort, aiming to evaluate their predictive value in a real-world setting objectively. In the ORIEN real-world dataset, we lacked specific tumor response information, i.e., complete response (CR)/partial response (PR)/stable disease (SD)/ progressive disease (PD); therefore, we utilized OS from the ICI medication start date as the main outcome for the prognostic evaluation. The Kaplan-Meier curves in Figure 3A compare the estimated OS probability associated with each of the 10 CEs identified in the ORIEN ICI cohort–across all patients who received ICIs. We understand that these patterns can be confounded by the cancer types and their distribution of CEs, but we still want to explore if well-established ecotypes, such as CE9 and CD10, can consistently exhibit similar effects at a pan-cancer level.

**Figure 3.**
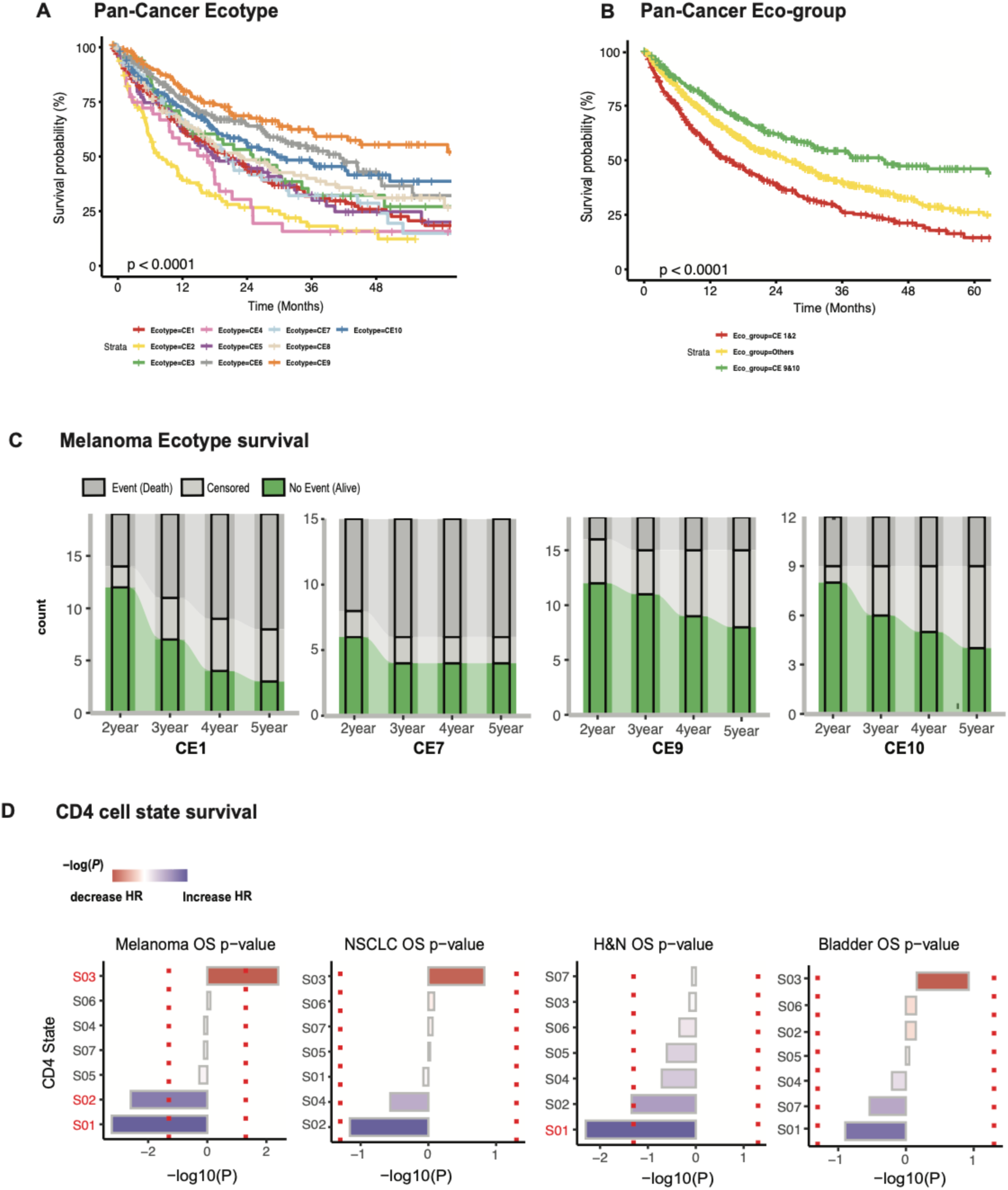
Associations Between Carcinoma Ecotypes, Cell States, and Survival Across Major Cancer Types. (**A**) Kaplan-Meier curves display patient overall survival across ten carcinoma ecotypes for overall survival (OS) within the ORIEN ICI cohort. (**B**) Kaplan-Meier survival curves with regrouping ten carcinoma eco-types (CE) into three sub-groups: CE1-CE2, CE9-CE10, and others (CE3-8). (**C**) Bar plots showing survival trends among melanoma patients over 2 to 5 years of treatment, categorized into CE1, CE7, CE9, and CE10 (**D**) Survival associations of CD4-specific cell states in melanoma, NSCLC, and H&N, and bladder with color coding for better survival outcomes (blue bar) or worse survival outcomes (red bar). The x-axis represents -log (p-value), the Y-axis characterizes cell state (S), and red dotted lines indicate a p-value of 0.05. Abbreviation: CE, carcinoma ecotype; H&N, head and neck cancer; HR, hazard ratio; NSCLC, non-small cell lung cancer; OS, overall survival.

Overall, CE9 exhibited the most favorable survival outcomes compared to all other ecotypes, closely followed by CE6 and CE10. Compared to the survival pattern observed in the TCGA discovery cohort, the favorable performance of CE6 is less expected compared to the two other proinflammatory CEs. Conversely, CE2, a lymphocyte deficiency CE, contributed to the least favorable patient survival outcome, followed by CE4 (myogenesis-associated CE). The remaining ecotype groups (CE1, CE3, CE5, CE7, and CE8) exhibited intermediate risk and did not show clear separations in their survival curves. The survival rates were further analyzed by grouping the ten identified ecotypes into three eco-groups: CE9 & CE10 (the proinflammatory group), CE1 & CE2 (the lymphocyte deficient group), and CE3-8 (Others) (Figure 3B). From the graph above, we can see the three eco-groups had clearly statistically significant separated survival curves (P<0.001, log-rank test); the CE9-10 group demonstrated the most favorable outcomes, while CE1-2 exhibited the least (Supplementary Figure 3). To better illustrate the ecotype-specific survival patterns, in Figure 3C, we additionally analyzed the population-level survival trajectories from two to five years for the four ecotypes (CE1, CE7, CE9, and CE10) that had the highest number of assigned patients among melanoma cases. As expected, there was a notable decrease in the number of patients experiencing no events (alive) over three years for CE1, while CE9, indicative of a favorable prognosis, shows a much smaller decrease. After that, we explored the prognostic implications of CD4 T CSs, which are crucial for immune regulation and the activation of cytotoxic T cells. It can be seen from the data in Figure 3D that CD4-S01 and CD4-S02 exhibited significant positive survival outcomes in melanoma and H&N, consistently showing beneficial effects in the other two cancer types as well. Finally, CD4-S03 (marked by gene *GLYCTK*) was associated with poorer survival outcomes in melanoma, NSCLC, and bladder cancer, although the mechanisms underlying CD4-S03 remain unclear. [Supplementary Figure 4C shows the CD4-S01 still exhibited significant positive association in both melanoma and HNC.]

### Multi-ecotype prognostic signature

To develop and optimize a melanoma-specific prognostic EcoTyper model using the ORIEN ICI melanoma data, we trained a multivariable Cox regression that ultimately selected five ecotypes: CE1, CE5, CE7, CE9, and CE10. The final melanoma-ICI patient risk score model was established as follows: 1.11 × CE1 + 1.49 × CE2 + 1.44 × CE3 - 2.12 × CE9 - 4.87 × CE10. Then, the predictive accuracy of this model was assessed using two external melanoma datasets: E1609 [a phase III adjuvant trial involving patients with resected cutaneous melanoma treated with ipilimumab (ipi) (3 or 10 mg/kg) or High-Dose Interferon Alfa-2b (HDI-α)] study^32^ and a harmonized ICI datasets^33–35^ comprising 334 patients with melanoma treated with ICIs. The prognostic value of the 5-CEs risk model across the entire E1609 melanoma cohort was presented in Figure 4A.

**Figure 4.**
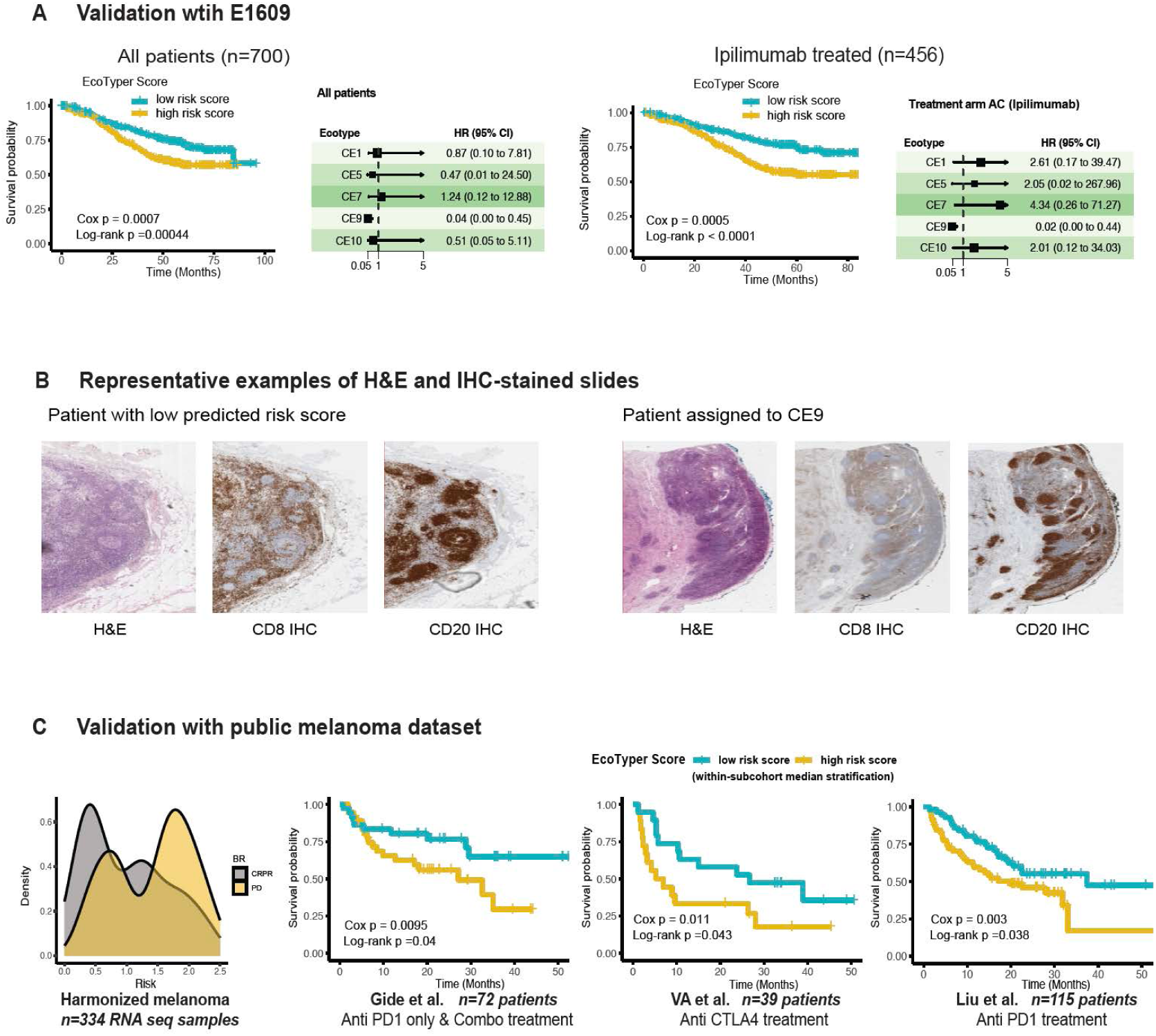
Validation of Predicted EcoTyper Risk Scores on External Melanoma Datasets. **(A)** Prognostic validation in the E1609 dataset: Kaplan-Meier overall survival curves compare patients categorized into low and high immuno-oncology (ICI) risk groups based on median scores with Cox regression and log-rank test p-values over 100 months of treatment. The forest plot describes eco-types associated with HR. Analyses were conducted on the entire E1609 cohort (left) and Ipilimumab-treated sub-arm (right). (**B**) Representative slides of melanoma tissue stained with H&E and IHC for markers CD8 and CD20, shown from left to right. Images represent a patient with a low predicted risk score and another assigned to CE9. **(C)** EcoTyper risk scores and prognosis were distributed in the harmonized melanoma dataset (n=334). Kaplan-Meier curves for overall survival over 50 months with log-rank p-values reported in individual datasets: Gide et al. (n=72), Van Allen et al. (n=39), and Liu et al. (n=115). Stratification is based on within-cohort median scores. Abbreviation: CE, carcinoma ecotype; CR/PR, complete response (CR)/partial response (PR); CTLA4, cytotoxic T-lymphocyte associated protein 4; H&E, Hematoxylin, and eosin stain; IHC, immunohistochemistry, OS, overall survival; PD, progressive disease, PD1, programmed cell death protein 1.

Stratification by the median of risk scores revealed that patients on either ipi or HDI-α with lower predicted scores exhibited significantly better survival outcomes than those in the high-risk score group (log-rank test, P<0.001). The same trend was observed in the subgroup of patients treated only with ipi, but it was not significant in the HDI-α treated cohort (non-ICI) (Supplementary Figure 5C). In the multivariable Cox regression setting, as illustrated in the forest plots, only CE9 remained significant in the all-patients cohort and ipi-treated cohort [Hazard ratio (HR) (95% CI); 0.04 (0.00 to 0.45), 0.02 (0.00 to 0.44), respectively] suggesting its potential value as a protective single-CE predictor.

An inspection of two melanoma tissue samples in the E1609 cohort was done by staining them with Hematoxylin and eosin stain (H&E) and immunohistochemistry (IHC), as presented in Figure 4B. One sample had a low melanoma-ICI patient risk score, and another one was classified under CE9. Notably, both samples displayed structures of lymphoid aggregation, which are frequently associated with better ICI outcomes, representing potential tertiary lymphoid structures, as indicated by the IHC results for CD8 (CD8+ T cells) and CD20 (B cells).

In the harmonized melanoma datasets^33–35^ (Figure 4C), the Progressive Disease (PD) cohort displayed a bimodal distribution, with the highest peak predominantly on the high-risk score side. The predictive accuracy of this ecotype risk score model was further validated using three external melanoma datasets.^33–35^

Within the cohort that received anti-PD-1 therapy from Gide *et al*.^33^, a marked survival difference was observed between the low and high-risk groups (p = 0.045). Besides, the VA *et al*.^35^ dataset, which included patients who received anti-CTLA4 therapy, echoed this finding, exhibiting distinct survival stratification (p = 0.011).

Furthermore, Liu *et al.*^34^ dataset involving patients on anti-PD1 therapy also showed the score’s ability to differentiate survival outcomes (p = 0.029), thus affirming the score’s prognostic potency. Further analysis of two alternative models; a full model with 8 CEs (Supplementary Figure 6A) and a single-CE model focused on CE9 (Supplementary Figure 7), confirmed that the selected 5-CE model consistently outperformed the others, suggesting its overall robustness.

### De novo ecotypes were discovered based on the ORIEN ICI dataset

As part of our exploratory analysis, we further performed the de novo discovery of CSs and ecotypes using the transcriptome data in the ORIEN ICI dataset. Based on CIBERSORTx, we first estimated the abundance of cell types and generated cell-type-specific gene expression profiles (GEPs) across nine cell types. Subsequently, employing the EcoTyper framework, we identified distinct transcriptional CSs within these profiles. Based on the ecotype discovery analysis pipeline, three distinct ecotypes were detected based on 27 different CSs. These newly identified ecotypes are labeled E1, E2, and E3, with 47, 32, and 39 RNAseq samples assigned to them, respectively (Figure 5A). Ecotype E1 was characterized by CD4-S04, which was related to gene CTLA4 and B-S01, CD8-S01, and others. On the contrary, Ecotype E2 consisted of various CSs, with S02 being the dominant one. Finally, Ecotype E3 was identified by the co-occurrence of CD8-S03 and EPI-S02, which was related to gene *SOX10* (Figure 5B).

**Figure 5.**
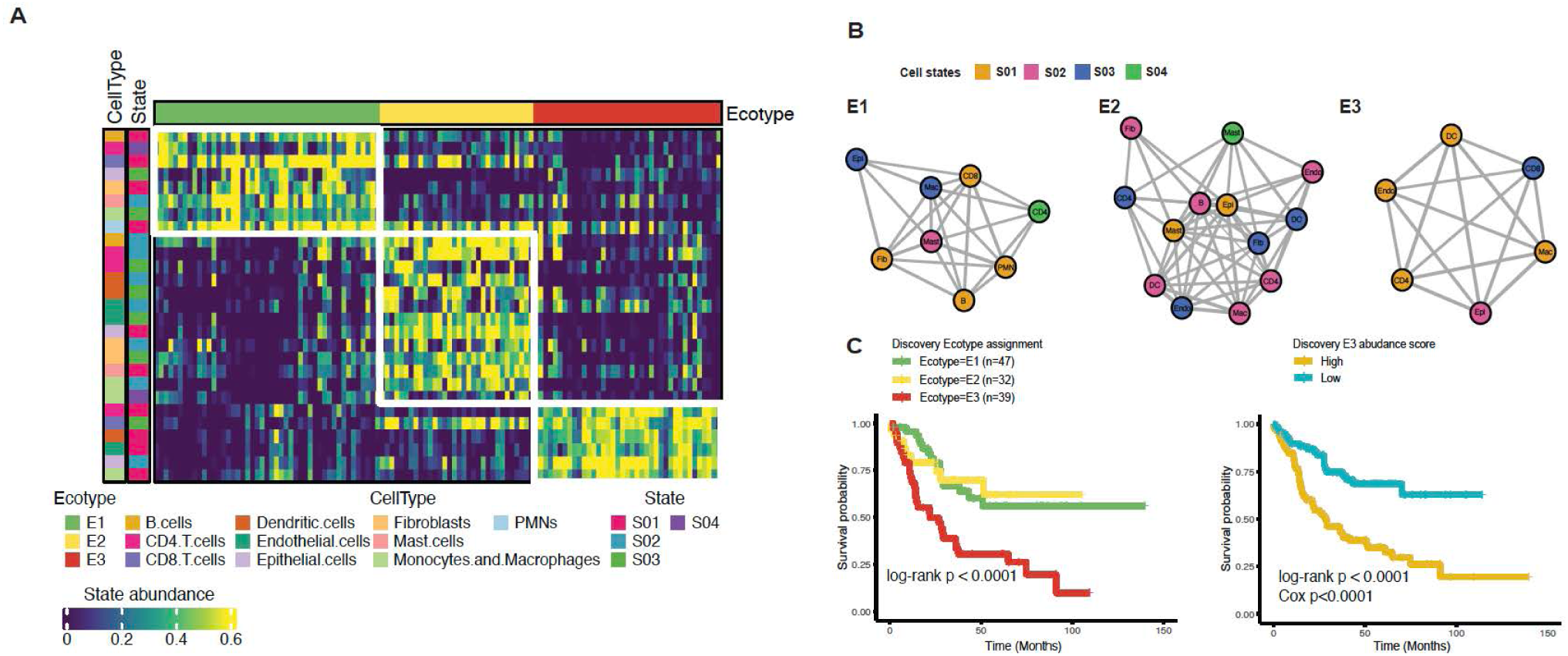
De-Novo Discovery and Characterization of Melanoma-Specific Cell States. **(A)** The heatmap displays the distribution of cell states and three de novo ecotypes (E1, E2, and E3) within the ORIEN melanoma ICI cohort. Each column in the heatmap corresponds to RNA sequencing samples, and each row shows the expression of cell state marker genes within a recovered ecotype. (**B**) Diagrams illustrate co-occurrence patterns between cell types and states within the three identified ecotypes. (**C**) Kaplan-Meier survival curves evaluating overall survival over 150 months of treatment among the three discovered ecotypes (on the left) with Log-rank p-value and for patients stratified into low and high abundance by median E3 abundance score (on the right) with Cox regression and log-rank test p-values—abbreviation: E, ecotype; S, cell state.

The survival analysis in Figure 5C indicates that patients categorized under Ecotype E3 experienced significantly poorer survival outcomes than those in E1 and E2 (log-rank test, P<0.0001). Moreover, the continuous score linked with E3 holds significant prognostic value; median-divided higher scores correspond to remarkably poorer survival (log-rank test, P<0.0001). This accentuates the crucial impact of Ecotype E3’s cellular attributes on patient prognosis, potentially attributed to its association with immune deficiency. Additionally, E1 and E2 show significant associations with survival outcomes. Supplementary Figure 8A indicates that higher abundance score groups in both are positively linked with better survival. We also investigated the links between the discovery Ecotypes and the recovery Ecotypes. E3 is predominantly associated with CE1, CE2, and CE7, which are the most immune-deficient groups (Supplementary Figure 8B). Furthermore, the distinct profile of Ecotype E3 could complement established prognostic ecotypes such as CE9, enhancing the profiling of immune interactions from derived real-world ICI data.

## DISCUSSION

In this study, we utilized the EcoTyper framework and ORIEN Avatar ICI cohort data to investigate how underlying immune CSs and tumor ecotypes influence response to immunotherapies. To the best of our knowledge, this is one of the largest studies examining the prognostic roles of carcinoma ecotypes in ICI treatment using real-world data. Public datasets, such as TCGA, used to train EcoTyper, were primarily selected for their high tumor purity and diverse cancer types.^36^ While these datasets are valuable, their generalizability is limited due to selective inclusion criteria and outdated treatment modalities. In the original publication of EcoTyper, the prognostic significance of ecotypes specific to ICI outcomes was validated on very limited datasets, such as IMvigor, a well-studied bladder cancer cohort receiving immunotherapy.^23^ We can overcome these limitations by leveraging the real-world ORIEN Avatar dataset, which features a much larger sample size and emphasizes high-quality patient data and clinical outcomes with modern treatments conducted under a common protocol at 18 leading cancer centers. This analysis enables a robust evaluation of the prognostic value of ecotypes in an independent real-world setting, providing more representative, clinically significant, and up-to-date insights into immunotherapy responses.

In the current study, the prognostic value of 10 ecotypes at the pan-cancer level was evaluated and subsequently categorized into three subgroups: an immune-deficient group (CE1 and CE2), a proinflammatory group (CE9 and CE10), and an intermediate-risk group (CE3-8). As expected, the immune-deficient group was associated with notably shorter survival, while the proinflammatory group demonstrated significantly favorable survival outcomes. These results demonstrated that major ecotype-associated prognostic signatures exist at a pan-cancer level regardless of histology. For cancer-specific optimization, tailored approaches are necessary to account for each cancer type’s unique TME and immune cell interactions.

We constructed a composite prognostic risk model in the melanoma cohort using ecotype abundance scores. Given that over 20% of patients were not assigned to specific ecotypes, our model incorporates all samples through continuous scores. This approach is particularly advantageous as it captures more information, even in ambiguous ecotypes like CE5 and CE8, which have comparable clustered CSs. Our model was designed to encompass the entire spectrum of ecotype variability, making it more effective in capturing immune CSs and their encapsulated prognostic significance. The final risk model selected CE1, CE5, CE7, CE9, and CE10. Among these, CE1 and CE7 were notably associated with TGF beta signaling. CE1 exhibited the strongest positive association with UV response downregulation and hypoxia, while CE9 demonstrated significant negative associations with these two hallmark pathways. Notably, these two ecotypes were also the ones to which most melanoma patients were assigned.

Then, we successfully validated the risk model for using an independent cohort derived from the E1609 phase III trial^32^ in melanoma and a harmonized cohort from three external independent studies^30,33,35^ of patients with melanoma, confirming that lower predicted risk scores are associated with improved survival and better response to immunotherapy. In samples with low predicted risk scores and patients assigned to CE9, potential lymphoid aggregation structures from H&E slides in the E1609 cohort were observed; this could indicate enrichment of immune-related functions at the spatial level. In the data with therapy response, patients with low predicted risk scores had a higher probability of achieving complete or partial response, whereas those with higher risk scores were more likely to experience progressive disease. Although CE9 appears to be a standalone prognostic marker, it did not consistently distinguish between low and high-risk groups in some external cohorts.

The predictive potential of major CSs and communities across all major cancer types in the ORIEN ICI cohort was also investigated. Unlike the extensively studied states and communities defined by CD8 T cells and B cells, which mostly have well-defined immune hot or cold states, CD4 T cells exhibit a broader range of CSs, with more than half of their states having unknown mechanisms. Through survival analysis for each state in each major ICI cancer type, we discovered that the CD4 T cell states S01 and S03 are generally the most consistent across cancers, with S01 associated with improved survival and S03 associated with poorer survival. We believe CD4 T cell states could be a promising complementary candidate to established ecotypes, warranting formal modeling and further analyses in the future. This analysis also tinted the need for more in-depth functional and translational research of CD4 T cells to fully understand their mechanisms and implications in cancer prognosis.

In addition to utilizing recovered CSs and ecotypes, we performed de novo discovery of CSs and ecotypes using the melanoma ORIEN ICI data and identified three melanoma-specific ecotypes: E1, E2, and E3. E1 was characterized by CD4-S04, associated with the immune checkpoint gene *CTLA4*, along with other CSs, including B-S01 and CD8-S01, and was linked to favorable survival outcomes. E3, identified by the co-occurrence of CD8-S03 and EPI-S02, was significantly associated with shorter survival. E2 consisted of a diverse mixture of CSs but did not show the same level of prognostic significance as E1 and E3. Overall, we could conclude that the distinct profile of E3 could complement established prognostic ecotypes such as CE9, enhancing the profiling of the immune functional status in the real-world setting.

This study has some limitations. First, the real-world nature of the ORIEN ICI cohort makes it challenging to obtain categorized RECIST responses and comprehensive details on all prior and ongoing treatments, which may impact our findings. Second, different therapy combinations, such as neoadjuvant chemotherapy regimens, may introduce bias and should be further explored in expanded cohorts.

Therefore, we relied on survival as the primary clinical outcome measure. Third, for calculating survival outcomes, we used the start date of the first ICI medication, which, for some patients, does not account for the duration or impact of prior treatments but is also consistent with how survival is computed in the context of clinical trials testing ICIs.

Additionally, our study relied primarily on established ecotypes due to the limited sample size required to discover and validate de novo ICI-specific cellular communities. However, the breadth and real-world origin of the ORIEN data provideharm a unique opportunity to thoroughly investigate the clinical utility of ecotypes that have not been explored in any existing datasets. In summary, our study comprehensively characterizes transcriptional CSs and cellular ecosystems within a real-world cohort treated with ICI. Utilizing the EcoTyper framework, we identified consistent ICI-specific prognostic TME ecotypes across diverse cancer types. We developed a composite ICI prognostic signature derived from five ecotypes, demonstrating robust predictive performance for survival outcomes among melanoma patients in both internal and external validation datasets.

In conclusion, our findings emphasize the potential of EcoTyper to facilitate personalized immunotherapy by refining patient selection and guiding therapeutic strategies that target distinct TME ecotypes. Future studies integrating EcoTyper with spatial omics CSs immune signatures could further refine its utility, ultimately contributing to the systematic and personalized application of immunotherapies and defining fundamental immune resistance mechanisms that guide future drug development.

## Acknowledgements

We are grateful to the participating patients and their family members as well as all research staff supporting the conduct of the Total Cancer Care protocol. Trial Registration: NCT03977402. This work was supported in part by ORIEN NOVA Team Science Award to AT; a NIH grant to X. Wang under award number R01DE030493; and the Biostatistics and Bioinformatics Shared Resource (BBSR) at the H. Lee Moffitt Cancer Center & Research Institute; an NCI designated Comprehensive Cancer Center (P30-CA076292).

## Institutional Review Board Statement

The study was conducted in accordance with the Declaration of Helsinki and approved by the Institutional Review Board (IRB; Pro00014441)

## Informed Consent Statement

Written informed consent was obtained at the participating institutions from all subjects involved in the original study protocol within which this project was nested.

## Data Availability Statement

All results relevant to this study are included in the article. Data are available upon reasonable requests to the corresponding author.

